# Serum but not mucosal antibody responses are associated with pre-existing SARS-CoV-2 spike cross-reactive CD4^+^ T cells following BNT162b2 vaccination in the elderly

**DOI:** 10.1101/2021.10.05.21264545

**Authors:** Lil Meyer-Arndt, Tatjana Schwarz, Lucie Loyal, Larissa Henze, Beate Kruse, Manuela Dingeldey, Kübrah Gürcan, Zehra Uyar-Aydin, Marcel A. Müller, Christian Drosten, Friedemann Paul, Leif E. Sander, Ilja Demuth, Roland Lauster, Claudia Giesecke-Thiel, Julian Braun, Victor M. Corman, Andreas Thiel

## Abstract

Advanced age is a main risk factor for severe COVID-19. However, low vaccination efficacy and accelerated waning immunity have been reported in this age group. To elucidate age-related differences in immunogenicity, we analysed human cellular, serological and salivary SARS-CoV-2 spike glycoprotein-specific immune responses to BNT162b2 COVID-19 vaccine in old (69-92 years) and middle-aged (24-57 years) vaccinees compared to natural infection (COVID-19 convalescents, 21-55 years). Serological humoral responses to vaccination exceeded those of convalescents but salivary anti-spike subunit 1 (S1) IgA and neutralizing capacity were less durable in vaccinees. In old vaccinees, we observed that pre-existing spike-specific CD4^+^ T cells are associated with efficient induction of anti-S1 IgG and neutralizing capacity in serum but not saliva. Our results suggest pre-existing SARS-CoV-2 cross-reactive CD4^+^ T cells as predictor of an efficient COVID-19 vaccine-induced humoral immune response in old individuals.

## Introduction

Global efforts have been mounted to develop efficient vaccines against coronavirus disease 2019 (COVID-19) (1). As severe COVID-19 mainly affects older individuals, many vaccination campaigns have prioritized the elderly population (2). However, vaccination efficacy is known to be decreased in this age group, particularly for primary vaccination (3). For COVID-19 vaccination - given the distinct homology of certain antigen target regions of severe acute respiratory syndrome coronavirus 2 (SARS-CoV-2) to human common cold coronaviruses (HCoV) - one possible explanation could be an age-related reduced number of pre-existing cross-reactive CD4^+^ T cells in old individuals (4, 5). To assess the immunogenicity of the COVID-19 vaccine in this particularly vulnerable age group and identify possible relations to pre-existing SARS-CoV-2-specific cross-reactivities, we examined systemic cellular and serological and salivary humoral SARS-CoV-2-specific immunity during the course of COVID-19 vaccination with BNT162b2 mRNA vaccine (Tozinameran™, Comirnaty™) in old and comorbid nursing home residents (n=18; mean age 83±6) and their middle-aged caregivers (n=14; mean age 47±10) at baseline (prior to first vaccination), at day 28 (d28, 7 days after second vaccination) and at day 49 (d49, 28 days after second vaccination). For comparison with naturally acquired immunity, we additionally analysed COVID-19 convalescents (of comparable age to the middle-aged cohort; mean age 36±11) after mild natural SARS-CoV-2 infection at ∼d28 (n=10), ∼d49 (n=16) or ∼d94 (n=11) after symptom onset.

## Materials and Methods

### Participants and ethics

The study was approved by the ethics committee of Charité – Universitätsmedizin Berlin (EA/152/20) and was conducted in accordance with the World Medical Association’s Declaration of Helsinki of 1964 and its later amendments. A written informed consent was obtained from all participants. The 39 participants (22 nursing home residents (old vaccinees), 17 caregivers (middle-aged vaccinees), all Caucasian) analysed for this study were recruited at three different nursing homes in Berlin between September and November 2020 and were available for follow-up visits 28 days and 49 days after their first COVID-19 vaccination in January and February 2021 (Table I). Furthermore, we collected saliva and blood samples of a total of 36 COVID-19 convalescents with mild disease course (World Health Organisation criteria for COVID-19 II) at ∼28 (n=10), ∼49 (n=16) or ∼94 days (n=11) post symptom onset. Baseline data of vaccinees and data of convalescents had been collected and partially analysed as part of the Charité Corona Cross (CCC) study (4). Visits included nasopharyngeal swabs, blood and saliva sampling at all time points. None of the participants took immunomodulating medication or reported immunocompromising comorbidities. 4 older and 3 middle-aged donors with signs of previous SARS-CoV-2 infection (either positive anti-S1 IgG levels or a S-I T cell stimulation index > 3.0 at baseline) were excluded from analysis.

**Table I.**
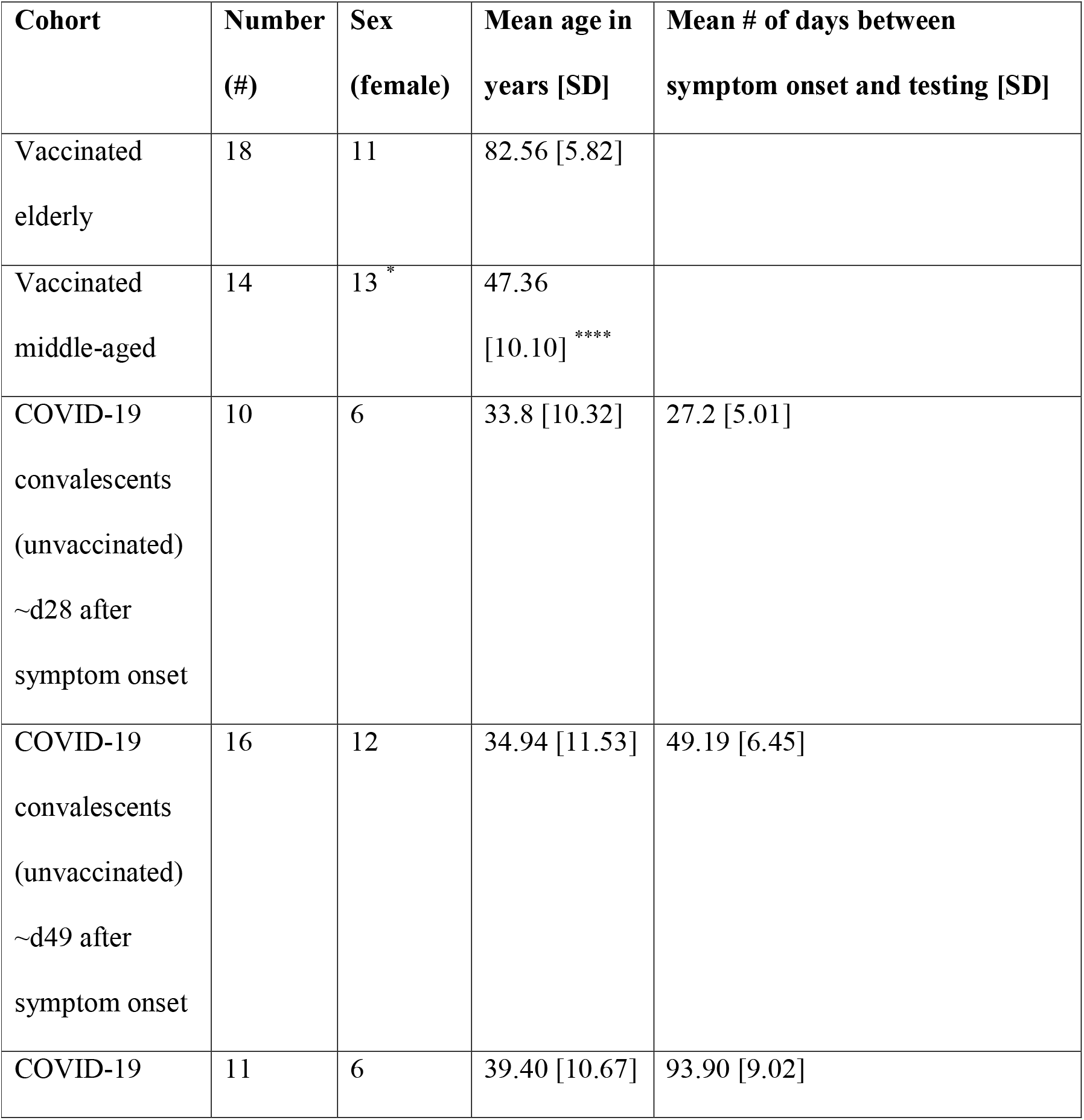
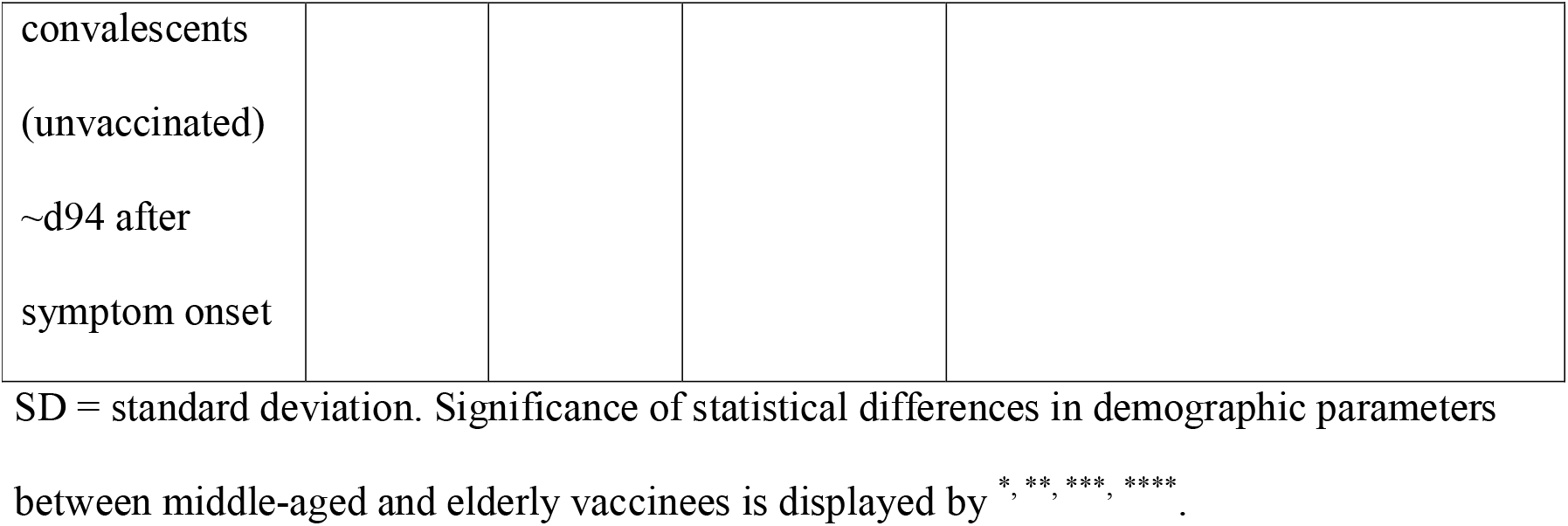
Donor characteristics.

### SARS-CoV-2 RT-PCR

Nasopharyngeal swabs were suspended in 4.3 ml Cobas PCR Media. RNA was extracted using the MagNA Pure 96 system (Roche). The viral RNA extraction was performed using 200 µl swab dilution eluted in 100 µl of extraction buffer. SARS-CoV-2 detection was based on two genomic targets (E- and N gene, TIB Molbiol) using 5 µl of the RNA eluate. Quantification of SARS-CoV-2 copy numbers was achieved using calibration curves with serial diluted photometrically quantified in-vitro transcribed RNA as described before (6). RT-PCR was performed using the LightCycler 480 II (Roche). Vaccinees who tested positive by RT-PCR in the study period (n=2) were excluded from all analyses.

### Blood sampling, serum preparation and PBMC isolation

Whole blood was collected in lithium heparin tubes for PBMC isolation and SST™II advance (all Vacutainer^®^, BD) tubes for serology. SST™II advance tubes were centrifuged at 1000 x g, 10 min and serum supernatant aliquots frozen at -80 °C until further use. PBMC were isolated by gradient density centrifugation according to the manufacturer’s instructions (Leucosep tubes, Greiner; Biocoll, Bio&SELL).

### Ex vivo T cell stimulation and flow cytometry

T cell in vitro stimulation and subsequent flow cytometric assessment of reactive CD4^+^ T cells was performed as described in detail previously (4). In brief, freshly isolated PBMC were stimulated with 11aa overlapping 15-mer PepMix™ SARS-CoV-2 spike glycoprotein peptide pool 1 or 2 (termed here S-I and S-II; JPT) or remained unstimulated and were subsequently incubated at 37 °C for 16 h. Surface staining was performed with the following fluorochrome-conjugated antibodies titrated to their optimal concentrations: CD3-FITC (REA613, Miltenyi), CD4-VioGreen (REA623, Miltenyi), CD8-VioBlue (REA734, Miltenyi), CD38-APC (REA671, Miltenyi), HLA-DR-PerCpVio700 (REA805, Miltenyi). Fixation and permeabilization were performed with eBioscience™ FoxP3 fixation and PermBuffer (Invitrogen). Intracellular staining was carried out for 30 min in the dark at room temperature with 4-1BB-PE (REA765, Miltenyi) and CD40L-PE-Vio770 (REA238, Miltenyi). All samples were measured on a MACSQuant^®^ Analyzer 16 (Miltenyi) according to the gating strategy illustrated in Supplemental Fig. 1.

### Anti-SARS-CoV-2 S1 ELISA in serum and saliva

Anti-SARS-CoV-2 spike glycoprotein subunit 1 (S1) IgG and IgA testing in sera was performed using a commercially available ELISA kit (Euroimmun) as previously described (7). Test results for sera were considered positive above an OD ratio (defined as absorbance difference between control and study sample) of 1.1 according to the manufacturer. The same ELISA kit was used for anti-SARS-CoV-2 IgA testing in saliva. The assay was performed with 1:100 diluted serum and 1:10 diluted saliva. Values were capped at an OD ratio of 10. Positivity thresholds have not yet been determined for saliva.

### Surrogate virus neutralization assay (sVNT)

A competition ELISA-based surrogate virus neutralization assay (sVNT; medac) mimicking the SARS-CoV-2 receptor (ACE2) binding process was used to identify neutralizing anti-SARS-CoV-2 antibodies in participant serum and saliva (8, 9). The assay was performed with 1:10 diluted serum and 1:5 diluted saliva respectively following the manufacturer’s instructions. Inhibition activity above 30% was considered positive in serum, no threshold has been defined for saliva yet.

### Data processing and statistical analysis

Study data were collected and managed using REDCap electronic data capture tools hosted at Charité (10). Flow cytometry data were analysed using FlowJo 10 (BD). Prism 9 (GraphPad) was used for data plotting. For statistical comparisons and correlation analyses, non-parametric testing (Mann-Whitney U test or Spearman regression including ROUT outlier tests) were performed. Mann-Whitney U tests were performed between corresponding timepoints if not indicated differently. Statistical significance was reported as follows: *p ≤ 0.05, **p ≤ 0.01, ***p ≤ 0.001. Correlation coefficients of Spearman correlations were reported as r. CD4^+^ T cell activation was plotted as stimulation index (Stimulation Index), i.e. frequency of CD40L^+^4-1BB^+^ CD4^+^ T cells in stimulated samples divided by unstimulated controls (zero background values were set to a minimum of 0.001).

## Results and Discussion

In this study, we investigated age-related differences in systemic and mucosal immune responses to COVID-19 mRNA vaccine BNT162b2 and compared to COVID-19 convalescents. First, we analysed anti-S1 IgG and anti-S1 IgA antibody levels and S1 neutralization capacity in serum as well as frequencies of peripheral antigen-reactive CD40L^+^ 4-1BB^+^ CD4^+^ T cells after *in vitro* stimulation with the N-terminal part (S1, covered by peptide mix S-I) and the C-terminal part (S2, peptide mix S-II) of the spike glycoprotein (Fig. 1a-e). In middle-aged donors, BNT162b2 vaccination induced a prompt and homogeneous response of anti-S1 IgG, anti-S1 IgA, S1-specific functional neutralization in serum and spike-reactive CD4^+^ T cells. In comparison, we observed significantly lower anti-S1 IgG and anti-S1 IgA levels and S1 neutralizing capacity in serum at both time points and lower T cell reactivity to S-I and S-II at ∼d49 in COVID-19 convalescents of comparable age after mild infection, which underlines the strong immunogenicity of BNT162b2 vaccine (Fig. 1a-e). However, in old vaccinees, particularly humoral vaccination responses were delayed and more heterogeneously distributed compared to the middle-aged cohort (Fig. 1a-e). For example, at d28, we did not detect anti-S1 IgG and anti-S1 IgA in five (28%) and eight (44%) old donors respectively (Fig. 1a and b; Supplemental Fig. 2) whereas all middle-aged donors exhibited strongly positive anti-S1 IgG levels. At d49, anti-S1 IgG and S1 neutralizing capacity (including one non-responder) were still significantly lower in the old vaccinees (Fig. 1a and c). Regarding the cellular response, all middle-aged and 89% (S-I) and 94% (S-II) respectively of the old donors acquired CD4^+^ T cell reactivity to S-I and S-II at d28. However, S-II-reactive T cell frequencies increased more homogenously and reached a higher level in the middle-aged than in the old cohort. At d49, both age groups reached comparable S-I- and S-II-reactive T cell levels (Fig. 1d-e). Consistently, reduced humoral and cellular vaccination responses in old individuals have been described for vaccines against influenza, yellow fever and tetanus as well as for COVID-19 (12-15).

**Fig. 1:**
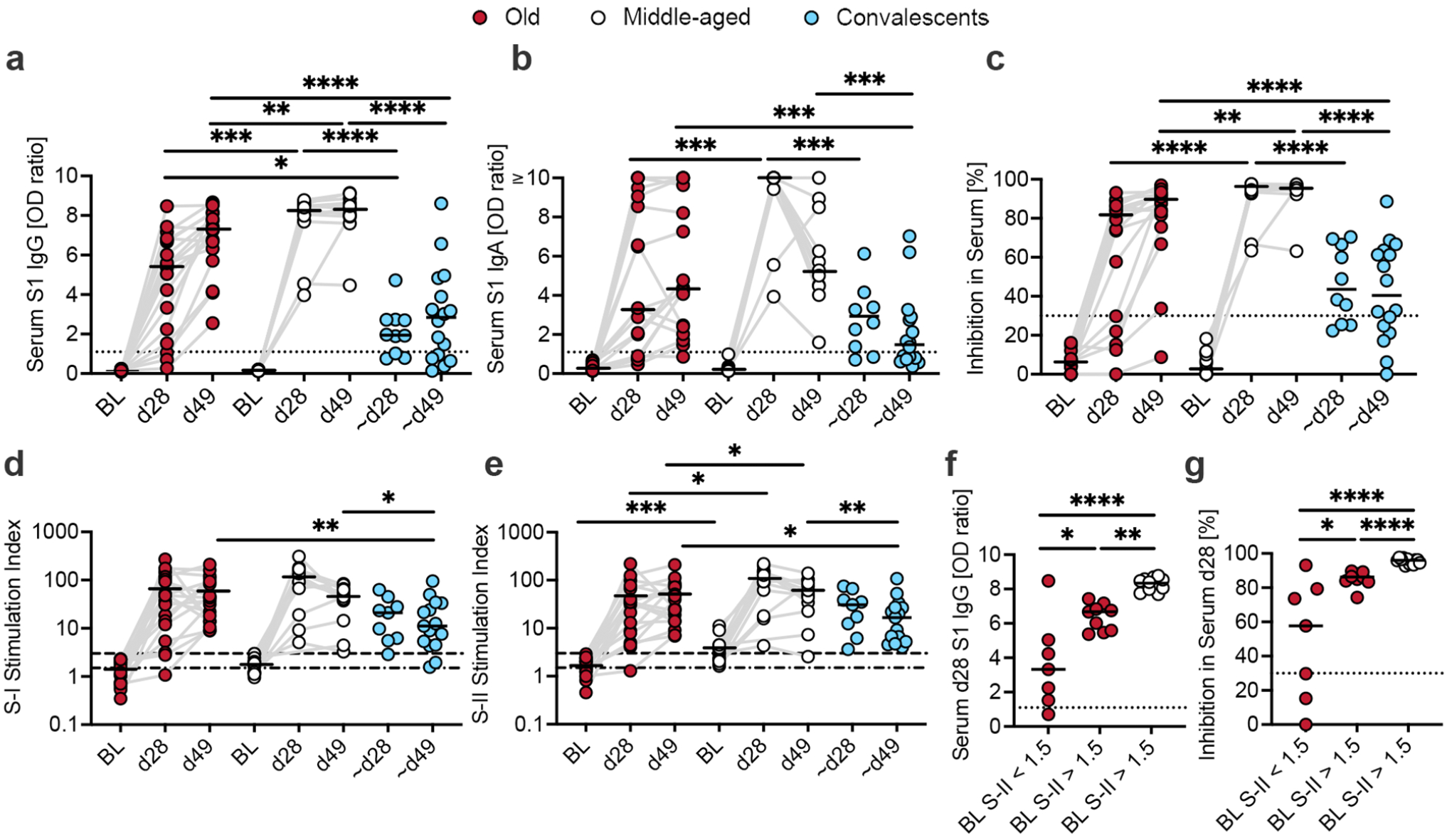
Serological antibody responses, neutralizing capacity and CD4^+^ T cell reactivity to S-I and S-II. **a** and **b**, anti-S1 serum IgG (**a**) and IgA (**b**) OD ratios in the old (red dots) and middle-aged (white) vaccinees at BL, d28 and d49 and in COVID-19 convalescents (blue) at ∼d28 or ∼d49. **c**, quantification of SARS-CoV-2 receptor (ACE2) binding inhibition in serum in percent in the old and middle-aged vaccinees at BL, d28 and d49 and in COVID-19 convalescents at ∼d28 or ∼d49. **d and e**, Stimulation Indices of S-I (**d**) and S-II (**e**) peptide pool-specific CD40L^+^ 4-1BB^+^ CD4^+^ T cells in the old and middle-aged vaccinees at baseline (BL), d28 and d49 and in COVID-19 convalescents at ∼d28 or ∼d49. **f and g**, anti-S1 serum IgG OD ratios (**f**) and inhibition in serum (**g**) in the old and middle-aged vaccinees at d28 grouped according to S-II-specific CD4^+^ T cell reactivity at BL (Stimulation Index > 1.5). Positivity thresholds: antibody OD ratio (dotted lines) > 1.1; neutralizing capacity (dotted lines) > 30%; CD4^+^ T cells Stimulation Index (dotdash lines) > 1.5 (positive, above background) and (dashed lines) > 3.0 (certainly positive; 6). Grey lines connect follow-up samples. p ≤ 0.05 = *, p ≤ 0.01=**, p ≤ 0.001=***, p ≤ 0.0001=**** according to the Mann-Whitney U test.

We have recently demonstrated that, in contrast to the N-terminal part, the C-terminal part of the spike glycoprotein contains highly conserved domains and triggers CD4^+^ T cell cross-reactivity to SARS-CoV-2 (11). Pre-existing T cell reactivity to S-II at baseline was significantly lower in old individuals than in the middle-aged cohort (Fig. 1e; Supplemental Fig. 3a). Possible explanations for this age-related reduction of cross-reactivity could be cellular senescence or impaired (oronasal) mucosal immunity (16-18). Remarkably though, in these old individuals, high levels of S-II-specific, but not S-I-specific, CD4^+^ T cells at baseline were associated with higher anti-S1 IgG and consistently with elevated S1 neutralizing capacity in serum at d28 (Fig. 1f-g; Supplemental Fig. 3b). In the light of the increased risk of the elderly for severe COVID-19 and the current discussions on their need of booster vaccinations, it is essential to identify and evaluate possible predictors of low vaccination efficiency particularly in this age group. Several studies have supported the notion of beneficial effects of pre-exposure SARS-CoV-2 cross-reactivity (4, 19, 20). However, whether this phenomenon has a direct effect on BNT162b2 immunogenicity in the elderly was unclear. Our findings here show that increased frequencies of pre-existing S-II-specific CD4^+^ T cells were associated with the efficiency of anti-S1 IgG and S1 neutralizing vaccination responses in the elderly. Cross-reactive CD4^+^ memory T cells expand faster upon antigen reactivation post-vaccination to aide B cell activation and class-switch and thus mount a more efficient antibody response.

We additionally investigated the presence of anti-S1 secretory IgA (sIgA) and S1 neutralization capacity in the saliva as a potential correlate of local mucosal protection from SARS-CoV-2 infection. We assume that the presence of S1-specific antibodies and S1 neutralizing capacity in the saliva may contribute to protection against SARS-CoV-2 infection and reduce local replication (21, 22). At d28 following vaccination, anti-S1 sIgA levels increased above their age groups’ maximum pre-vaccination level in all middle-aged (0.45 OD ratio) but only 60% of the old donors (0.31 OD ratio; Fig. 2a). Consistently, an increase in S1 neutralizing activity in saliva was detected in most vaccinees at d28 (Fig. 2b). This response, however, was transient and anti-S1 sIgA and salivary S1 neutralization dropped to pre-vaccination levels in all vaccinees within four weeks (d49) after the second vaccination (Fig. 2a and b). There was no correlation between anti-S1 sIgA or salivary S1 neutralizing capacity and pre-existing cross-reactive CD4^+^ T cells (Fig. 2c and d). Intriguingly, compared to aged-matched middle-aged vaccinees, COVID-19 convalescents exhibited significantly higher anti-S1 sIgA levels and S1 neutralizing capacity in the saliva at ∼d49 after symptom onset (Fig. 2a and b). Unlike salivary S1 neutralizing capacity, anti-S1 sIgA remained significantly increased in convalescents at ∼d94 indicating that anti-S1 sIgA does not correspond to neutralizing activity following infection at later time points. However, we found a correlation between anti-S1 sIgA levels and salivary S1 neutralization in convalescents at ∼d28 and at ∼d49, which was not observed in vaccinees (Supplemental Fig. 4). This suggests that neutralizing capacity in the saliva following vaccination may not only rely on anti-S1 sIgA but possibly anti-S1 IgG, which is consistent with reports on detectable anti-S1 IgG in the saliva of vaccinated individuals (23). In COVID-19 convalescents, anti-S1 sIgA secretion in salivary glands (and salivary S1 neutralizing activity) is likely induced by locally primed B and T cells in nasopharyngeal lymph nodes and/or tonsils (24). This is underlined by an increase in salivary neutralization capacity and sIgA in convalescents from d28 to d49 after infection (Fig. 2a and b) and may indicate generation of tissue-resident plasma cells after mucosal priming. In contrast, the more transient presence of anti-S1 sIgA in the saliva of vaccinated individuals could be the result of transfusion of serum-derived anti-S1 IgA through the endothelium into the oral mucosa (25). Currently, vaccines for intranasal application are in development, which may fill the gap in mucosal immunity observed here (26).

**Fig. 2:**
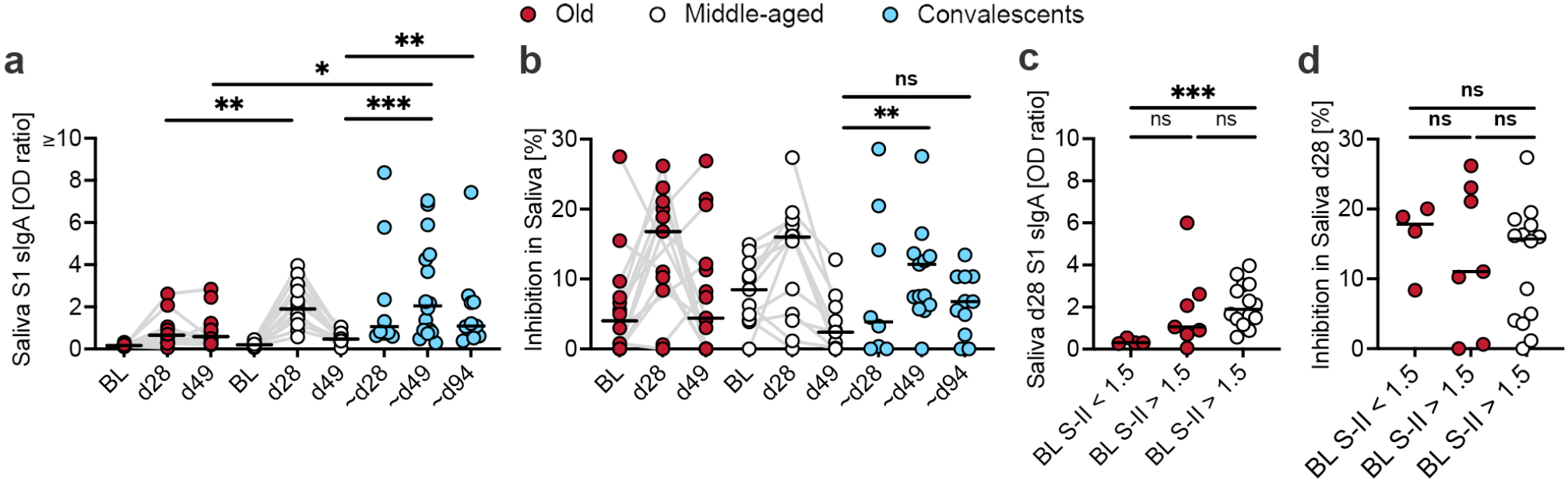
Salivary immune responses. **a**, anti-S1 saliva sIgA OD ratios in the old (red dots) and middle-aged (white) vaccinees at BL, d28 and d49 and in COVID-19 convalescents (blue) at ∼d28, ∼d49 or ∼d94. **b**, quantification of SARS-CoV-2 receptor (ACE2) binding inhibition in saliva in percent in the old and middle-aged vaccinees at BL, d28 and d49 and in COVID-19 convalescents at ∼d28, ∼d49 or ∼d94. **c and d**, anti-S1 saliva sIgA OD ratios (**c**) and inhibition in saliva (**d**) in the old and middle-aged vaccinees at d28 grouped according to S-II-specific CD4^+^ T cell reactivity at BL (Stimulation Index > 1.5). Salivary anti-S1 sIgA and ACE2 binding inhibition are displayed as means of up to three saliva samples on consecutive days (one per day for three days). Grey lines connect follow-up samples. ns = non-significant, p ≤ 0.05 = *, p ≤ 0.01=**, p ≤ 0.001=*** according to the Mann-Whitney U-test.

Taken together, our findings indicate that the presence of anti-S1 sIgA and S1 neutralizing capacity in the saliva after vaccination is of shorter duration and lower magnitude than after natural infection highlighting the need to determine the role of mucosal immunity, e.g., in the form of sIgA in saliva, for evaluation of SARS-CoV-2 immunity and its transmission. Furthermore, we demonstrate that BNT162b2 induces strong immune responses in middle-aged as well as most old and comorbid individuals. However, for some old individuals, the serological response to vaccination is hampered and may leave these individuals at higher risk of infection and severe disease courses, thus promoting recommendations for regular immune status check-ups and further vaccination boosts. Importantly, we show here that pre-existing SARS-CoV-2 spike glycoprotein cross-reactive memory T cells are associated with vaccination efficiency in the elderly and may generally contribute to the high responsiveness to COVID-19 vaccines.

## Data Availability

The datasets generated during the current study are available from the corresponding author on reasonable request.

## Acknowledgments and Funding

We thank the CCC Study Group.

## Abbreviations

COVID-19: coronavirus disease 2019
d28: day 28 after first vaccination
d49: day 49 after first vaccination
∼d28: around day 28 after COVID-19 symptom onset
∼d49: around day 49 after COVID-19 symptom onset
∼d94: around day 94 after COVID-19 symptom onset
SARS-CoV-2: severe acute respiratory syndrome coronavirus 2
sIgA: secretory IgA
sVNT: surrogate virus neutralization assay
S1: SARS-CoV-2 spike glycoprotein subunit 1
S-I: peptide mix representing the SARS-CoV-2 spike glycoprotein N-terminal part
S-II: peptide mix representing the SARS-CoV-2 spike glycoprotein C-terminal part mix

