## Supplemental Data for "Serum but not mucosal antibody responses are associated with pre-existing SARS-CoV-2 spike cross-reactive CD4^+^ T cells following BNT162b2 vaccination in the elderly"

### Supplemental Figure

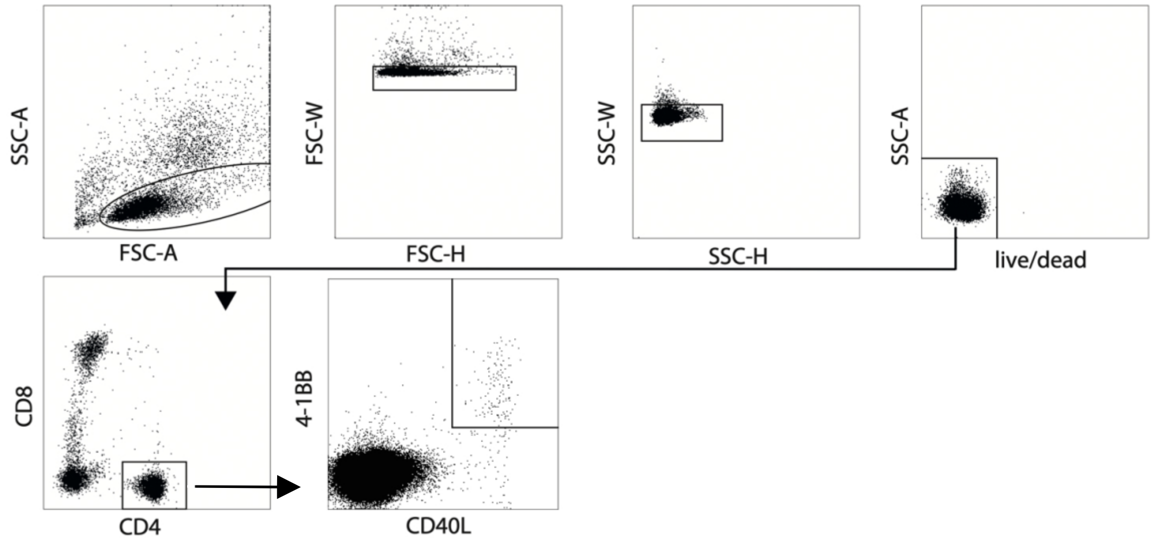


**Supplemental Fig. 1: Gating scheme.** The gating scheme is displayed for one representative donor's CEFX-stimulation utilized as positive control. Doublets were removed from lymphocytic populations via FSC-H vs -W and SSC-H vs -W and dead cells via Zombie Yellow live/dead stain. Subsequently, $CD4^+$ cells were gated within $CD3^+$ T cells. Antigen-reactive $CD4^+$ T cells were identified via CD40L and 4-1BB staining.

### Supplemental Figure 2

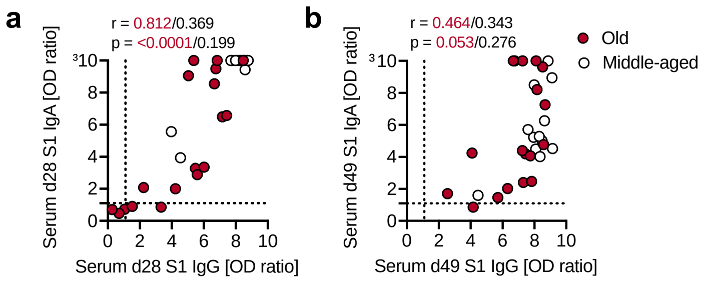


**Supplemental Fig. 2: Correlation of anti-S1 IgG and anti-S1 IgA in serum after vaccination.** Anti-S1 IgG OD ratios in serum correlated with anti-S1 IgA in serum at d28 (**a**) or d49 (**b**), respectively, in the old (red dots) and middle-aged (white dots) vaccinees. Positivity threshold (dotted lines) > 1.1. r = correlation coefficient, $p \leq 0.05$ = significant according to Spearman's rank.

### Supplemental Figure 3

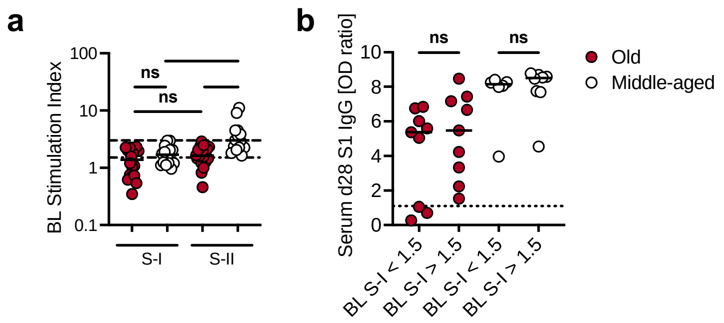


**Supplemental Fig. 3: Pre-existing CD4$^+$ T cell reactivity to S-I and S-II. a,** Stimulation Index of S-I- and S-II-specific CD40L$^+$ 4-1BB$^+$ CD4$^+$ T cells in the very old and middle-aged vaccinees at BL grouped according to stimulation for direct comparison. **b**, anti-S1 serum IgG OD ratios in the old and middle-aged vaccinees at d28 grouped according to S-I-specific CD4$^+$ T cell reactivity at BL (Stimulation Index > 1.5). Positivity threshold serology (dotted lines) > 1.1; CD4$^+$ T cells Stimulation Index (dotdash lines) > 1.5 (positive, above background) and (dashed lines) > 3.0 (certainly positive; 6). ns = non-significant, $p \leq 0.05$ = *, $p \leq 0.01$=**, $p \leq 0.001$=*** according to the Mann-Whitney U-test.

### Supplemental Figure 4

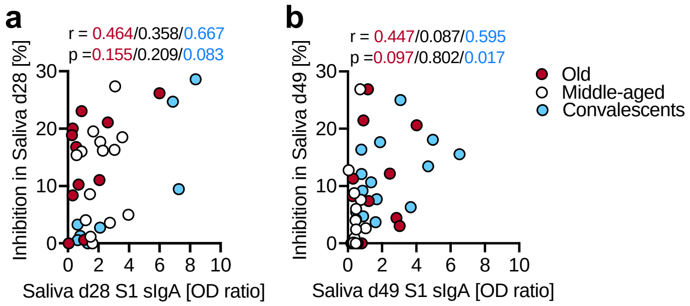


**Supplemental Fig. 4: Correlation of virus inhibition with anti-S1 sIgA in saliva after vaccination or infection.** S1 virus inhibition in saliva at d28 (**a**) or d49 (**b**) correlated with salivary anti-S1 sIgA OD ratios at d28 (**a**) or d49 (**b**) respectively in the old (red dots) and middle-aged (white dots) vaccinees and in convalescents (blue dots). r = correlation coefficient, $p \leq 0.05$ = significant according to Spearman's rank.
